# Prevalence and Correlates of HIV Among Children Born to HIV-Negative Mothers in Zambia: A Secondary Analysis of the 2024 Zambia Demographic and Health Survey

**DOI:** 10.64898/2026.03.19.26348774

**Authors:** Chibesa Chalwe, Picket Munkombwe, Beatrice Mulenga

## Abstract

Despite substantial progress in preventing mother-to-child transmission (PMTCT) of HIV in Zambia, HIV infection among children whose mothers test HIV-negative represents an understudied phenomenon that may reflect non-vertical transmission pathways. This cross-sectional secondary analysis of the 2024 Zambia Demographic and Health Survey (ZDHS) examined the prevalence and correlates of HIV among children aged 2–14 years whose mothers tested HIV-negative. Children with valid HIV blood test results were linked to their co-resident mothers’ HIV test results using household-level identifiers. HIV prevalence with 95% confidence intervals was estimated by age group, sex, residence, province, and wealth quintile. Bivariate associations were assessed using chi-square tests and odds ratios. Among 13,960 children of HIV-negative mothers, 69 (0.49%; 95% CI: 0.39%–0.63%; weighted: 0.58%) tested HIV-positive. Prevalence varied substantially by province (*p* < 0.001), with Copperbelt Province exhibiting the highest burden at 2.97%. Urban children had significantly higher prevalence than rural children (1.10% vs. 0.26%; OR = 3.68, 95% CI: 2.25–6.03, *p* < 0.001). A paradoxical wealth gradient was observed, with children in richer households showing higher prevalence than those in the poorest households (OR = 4.93, 95% CI: 2.19–11.10, *p* < 0.001). Maternal education was marginally associated with child HIV status (*p* = 0.04). Neither child sex (*p* = 0.10) nor age group (*p* = 0.29) was significantly associated with HIV positivity. A substantial proportion of HIV-positive children in Zambia have HIV-negative mothers, highlighting important gaps in the current PMTCT-focused paradigm and calling for expanded prevention strategies that address transmission risks beyond vertical transmission.

## Introduction

Zambia continues to bear one of the highest HIV burdens in sub-Saharan Africa. According to the 2024 Zambia Demographic and Health Survey (ZDHS), HIV prevalence among adults aged 15–49 years was 8.7%, with marked geographic heterogeneity across the country’s ten provinces [1]. This represents a notable decline from the 11.1% prevalence reported among adults aged 15–49 in the 2018 ZDHS [2] and the 11.0% prevalence documented by the 2021 Zambia Population-based HIV Impact Assessment (ZAMPHIA) among adults aged 15 years and older [3]. Despite this downward trajectory, the absolute number of people living with HIV remains substantial, and the epidemic continues to disproportionately affect women, urban populations, and specific provinces [1,4]. Importantly, the 2024 ZDHS also documented an HIV prevalence of 1.2% among children aged 2–14 years [1], confirming that paediatric HIV remains a persistent public health challenge even as adult prevalence declines.

Zambia has made considerable strides in reducing mother-to-child transmission (MTCT) of HIV. The national MTCT rate declined from approximately 30% in 2005 to 5.9% by 2025, reflecting the successful scale-up of prevention of mother-to-child transmission (PMTCT) services, including the adoption of Option B+ (lifelong antiretroviral therapy for all pregnant and breastfeeding women living with HIV) and expanded early infant diagnosis [5,6]. Nationally, new HIV infections fell from 67,585 in 2005 to an estimated 29,782 in 2025, with an estimated 3,000 children aged 0–2 years newly infected through MTCT in the most recent reporting period [5,7]. However, the national MTCT rate remains above the global elimination target of below 5%, and notably, Copperbelt Province recorded the highest number of new HIV infections in 2024, underscoring persistent subnational disparities [5].

Against this backdrop, a less well-characterised dimension of the paediatric HIV epidemic concerns children who test HIV-positive despite being born to mothers who test HIV-negative. Such children represent a population for whom classical vertical transmission is an unlikely or incomplete explanation for their HIV status. Several mechanisms may account for HIV acquisition in this group: healthcare-associated transmission through contaminated medical equipment, unsafe injections, or inadequately screened blood products; and, particularly among older children, sexual abuse or early sexual debut [8,9]. Understanding the magnitude and distribution of HIV in this specific subpopulation is essential for identifying programmatic gaps and tailoring interventions beyond the conventional PMTCT cascade.

The Demographic and Health Survey (DHS) programme provides a unique opportunity to examine this question. The DHS conducts population-based HIV testing of both women and children within the same household, allowing researchers to link maternal and child HIV status and to identify children whose seropositivity cannot be straightforwardly attributed to vertical transmission [1]. The 2024 ZDHS—the seventh DHS conducted in Zambia since 1992—collected data from 17 January to 7 July 2024 and tested women aged 15–49, men aged 15–59, and children aged 2–14 for HIV using both rapid diagnostic tests and laboratory-based dried blood spot (DBS) analysis [1]. Prior analyses using DHS data from other countries in the region, including Mozambique, Malawi, and Zimbabwe, have documented small but non-negligible proportions of HIV-positive children born to HIV-negative mothers, pointing to the existence of under-recognised transmission pathways [10,11].

To date, however, there has been limited investigation of this phenomenon using the most recent Zambian data. The 2024 ZDHS provides the most current nationally representative dataset with linked maternal and child HIV test results, enabling a timely assessment of the prevalence and sociodemographic correlates of HIV among children born to HIV-negative mothers. Such an analysis can inform targeted prevention efforts and contribute to the evidence base on non-MTCT paediatric HIV acquisition in high-burden settings. This is particularly salient given recent reductions in international funding for HIV programmes in Zambia—including a US$367 million reduction in the 2025 PEPFAR budget—which necessitate more efficient, evidence-based targeting of prevention resources [7].

This study aimed to: (1) quantify the burden of HIV among children aged 2–14 years whose mothers tested HIV-negative in the ZDHS 2024; (2) examine age-specific patterns of paediatric HIV in this population; and (3) identify sociodemographic factors associated with HIV positivity among these children.

## Materials and methods

### Study design

This was a cross-sectional secondary analysis of the 2024 Zambia Demographic and Health Survey (ZDHS 2024). The ZDHS is a nationally representative household survey conducted by the Zambia Statistics Agency (ZamStats) in partnership with the Ministry of Health, the University Teaching Hospital Virology Laboratory, and the Department of Demography, Population Sciences, Monitoring and Evaluation at the University of Zambia, with technical assistance from ICF through The DHS Program [1].

### Ethics statement

This study used publicly available, de-identified DHS data accessible through The DHS Program data repository (https://dhsprogram.com). The datasets (HIV Test Recode ZMAR81FL and Person Recode ZMPR81FL) were accessed on 1 March 2026 following registration and approval of a data access request. The original ZDHS 2024 received ethical approval from the Zambia National Health Research Authority (NHRA) and the ICF Institutional Review Board (IRB). During the original survey, informed consent was obtained from all adult participants prior to data and specimen collection. For children aged 2–9 years, parental or responsible adult consent was obtained. For children aged 10–14 years, both minor assent and parental consent were required. For adolescents aged 15–17 years, both adolescent assent and parental consent were required [1]. At no point during or after data access did the authors have access to information that could identify individual participants. All datasets obtained from The DHS Program are stripped of personally identifiable information prior to public release, and no attempt was made to identify any individual participant. No additional ethical approval was required for this secondary analysis of anonymised data.

### Sampling design

The survey employed a two-stage stratified cluster sampling design. The sampling frame was based on the 2022 Census of Population and Housing of the Republic of Zambia. Zambia is administratively divided into 10 provinces, further subdivided into 116 districts, 156 constituencies, and 1,858 wards. Enumeration areas from the 2022 census served as primary sampling units. A total of 545 clusters were selected, with a fixed number of 25 households selected per cluster, yielding a total sample of 13,625 households. Data collection was carried out from 17 January to 7 July 2024 by 22 field teams [1].

### Study population

The study population comprised children aged 2–14 years who met the following inclusion criteria: (1) blood specimens were collected and tested for HIV as part of the ZDHS HIV testing component; (2) HIV test results were valid (HIV-positive or HIV-negative); and (3) children were linked to their mothers in the same household, where the mother also had a valid HIV test result classified as HIV-negative.

### Variables

The primary outcome was the child’s HIV status, classified as positive or negative based on the DHS HIV testing protocol. The DHS uses a standardised laboratory algorithm for HIV testing of blood samples collected via dried blood spots (DBS), with confirmatory testing for positive results [1].

Independent variables were selected a priori based on the existing literature on paediatric HIV epidemiology and data availability within the DHS. These included: province of residence (ten provinces), type of place of residence (urban vs. rural), child’s sex (male vs. female), child’s age group (2–4, 5–9, and 10–14 years), household wealth index quintile (poorest, poorer, middle, richer, richest), and mother’s highest educational attainment (no education, primary, secondary, higher).

### Statistical analysis

All analyses were conducted using survey-weighted estimates to account for the complex sampling design of the DHS, including stratification, clustering, and unequal probability of selection. Sampling weights provided in the DHS dataset were applied throughout.

Weighted HIV prevalence was estimated overall and stratified by each independent variable. Bivariate associations between each sociodemographic factor and child HIV status were assessed using design-adjusted Rao-Scott chi-square tests. Crude odds ratios (ORs) with 95% confidence intervals (CIs) were calculated for binary and ordinal variables, with reference categories chosen as the lowest-risk group (e.g., rural residence, poorest wealth quintile). Statistical significance was defined as *p* < 0.05.

All analyses were performed in Python (version 3.14) using the pandas, NumPy, and statsmodels libraries, with survey-weighted estimates computed using the linearmodels package.

## Results

### Study population and participant flow

The participant flow from the ZDHS 2024 HIV Test Recode through to the final analytical sample is shown in Fig 1. The dataset contained 48,444 records, of which 22,089 were children aged 2–14 years. Of these, 22,042 (99.8%) had valid HIV test results, while 47 had inconclusive results and were excluded. Of the 22,042 children with valid results, 17,164 (77.9%) had mothers identified as residing in the same household. Among these, 15,956 mothers had valid HIV test results: 13,983 (87.6%) tested HIV-negative, 1,719 (10.8%) tested HIV-positive, and 254 (1.6%) had inconclusive or HIV-2 positive results. After excluding children whose mothers had inconclusive results and 23 records with discrepant age data, the final analytical sample comprised 13,960 children of HIV-negative mothers.

**Fig 1.**
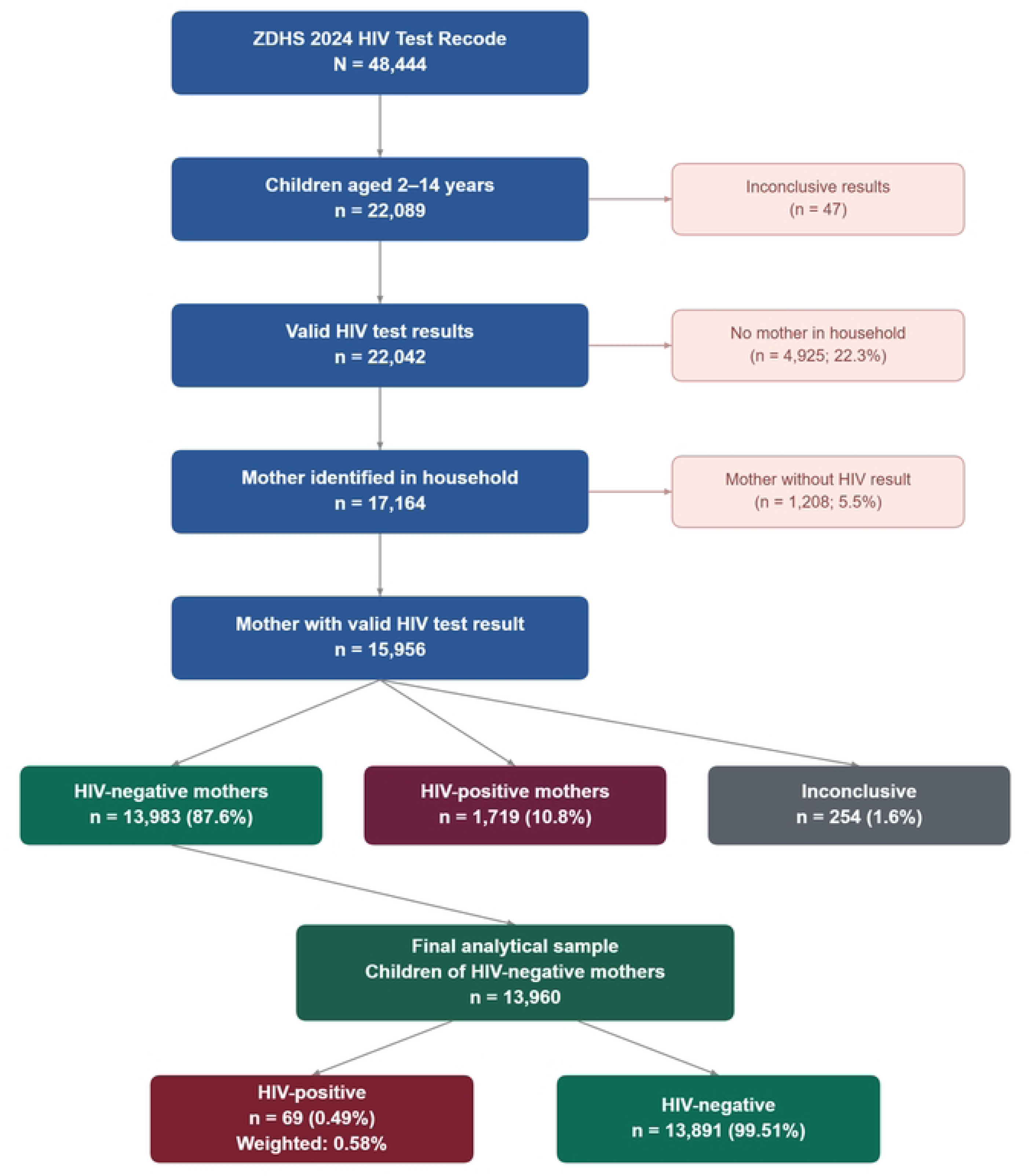
Participant flow diagram. Flow of participants from the ZDHS 2024 HIV Test Recode dataset to the final analytical sample of 13,960 children aged 2–14 years of HIV-negative mothers.

### Overall HIV prevalence

Among the 13,960 eligible children, 69 tested positive for HIV, yielding a weighted prevalence of 0.58%.

### HIV prevalence by sociodemographic characteristics

Table 1 presents the HIV prevalence among children of HIV-negative mothers stratified by sociodemographic characteristics.

**Table 1.** Summary of bivariate associations with HIV positivity among children aged 2–14 years born to HIV-negative mothers, 2024 ZDHS.

| Variable | Key OR (95% CI) | *p*-value |
| --- | --- | --- |
| Province | — | < 0.001 |
| Residence (urban vs. rural) | 3.68 (2.25–6.03) | < 0.001 |
| Wealth quintile (poorest to richest) | 4.93 (2.19–11.10)<sup>a</sup> | < 0.001 |
| Mother’s education (none to higher) | — | 0.04 |
| Child sex (male vs. female) | 1.54 (0.95–2.50) | 0.10 |
| Child age group (2–4 / 5–9 / 10–14) | — | 0.29 |

OR = odds ratio; CI = confidence interval. <sup>a</sup>Richer vs. poorest quintile

### Prevalence by province

HIV prevalence varied markedly across Zambia’s ten provinces (Table 1; *p* < 0.001). Copperbelt Province exhibited the highest weighted prevalence at 2.97% (34 of 1,220 children), approximately five times the national average (Fig 2).

**Fig 2.**
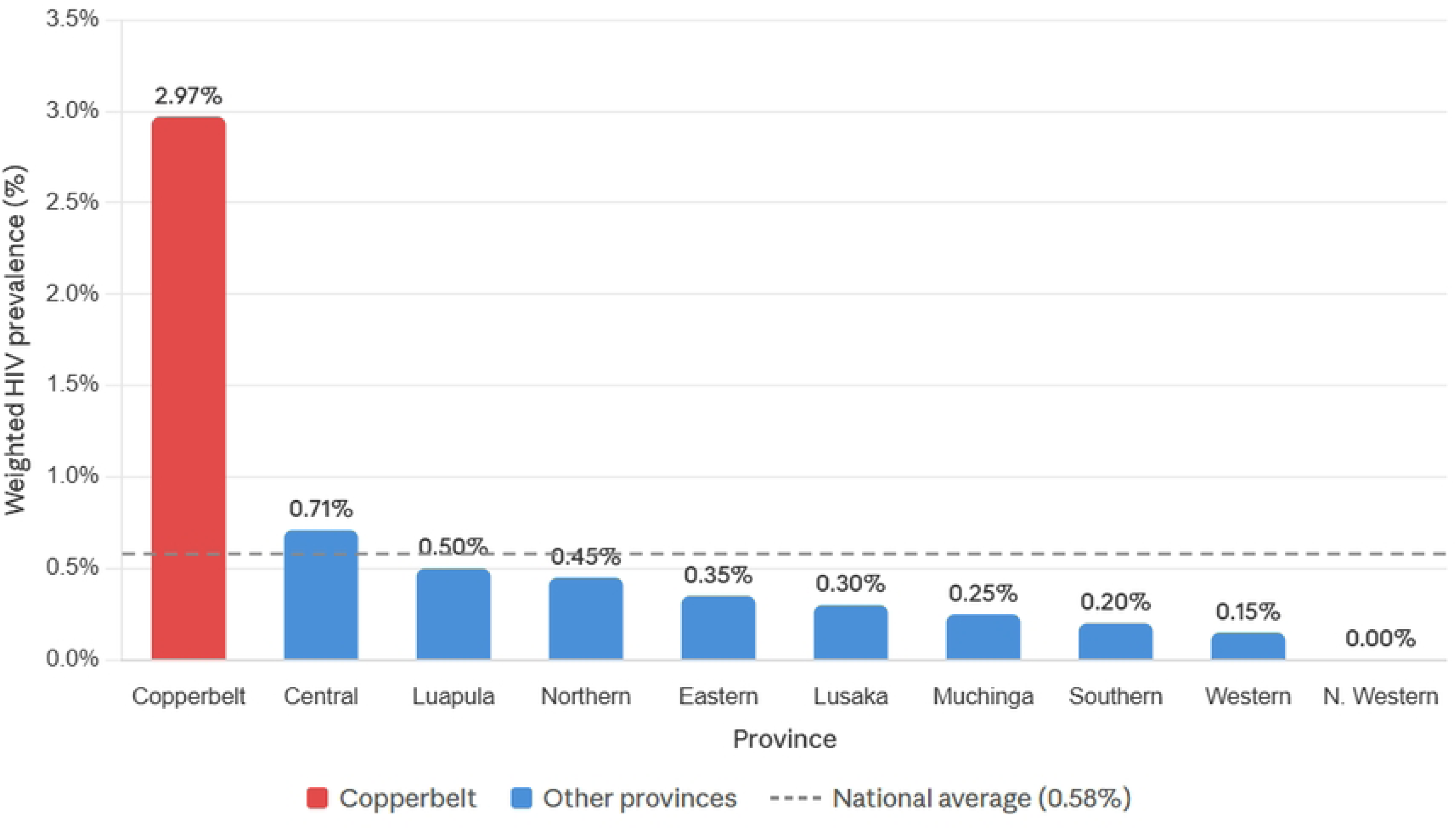
HIV prevalence by province among children of HIV-negative mothers, ZDHS 2024. Copperbelt Province had the highest burden at 2.97%. Dashed line indicates the national weighted average (0.58%).

### Prevalence by type of residence

Children residing in urban areas had a weighted HIV prevalence of 1.10% (44 of 4,535), compared with 0.26% (25 of 9,425) among rural children (Fig 3B). The odds of HIV positivity were approximately 3.7 times higher in urban than in rural areas (OR = 3.68, 95% CI: 2.25–6.03, *p* < 0.001).

**Fig 3.**
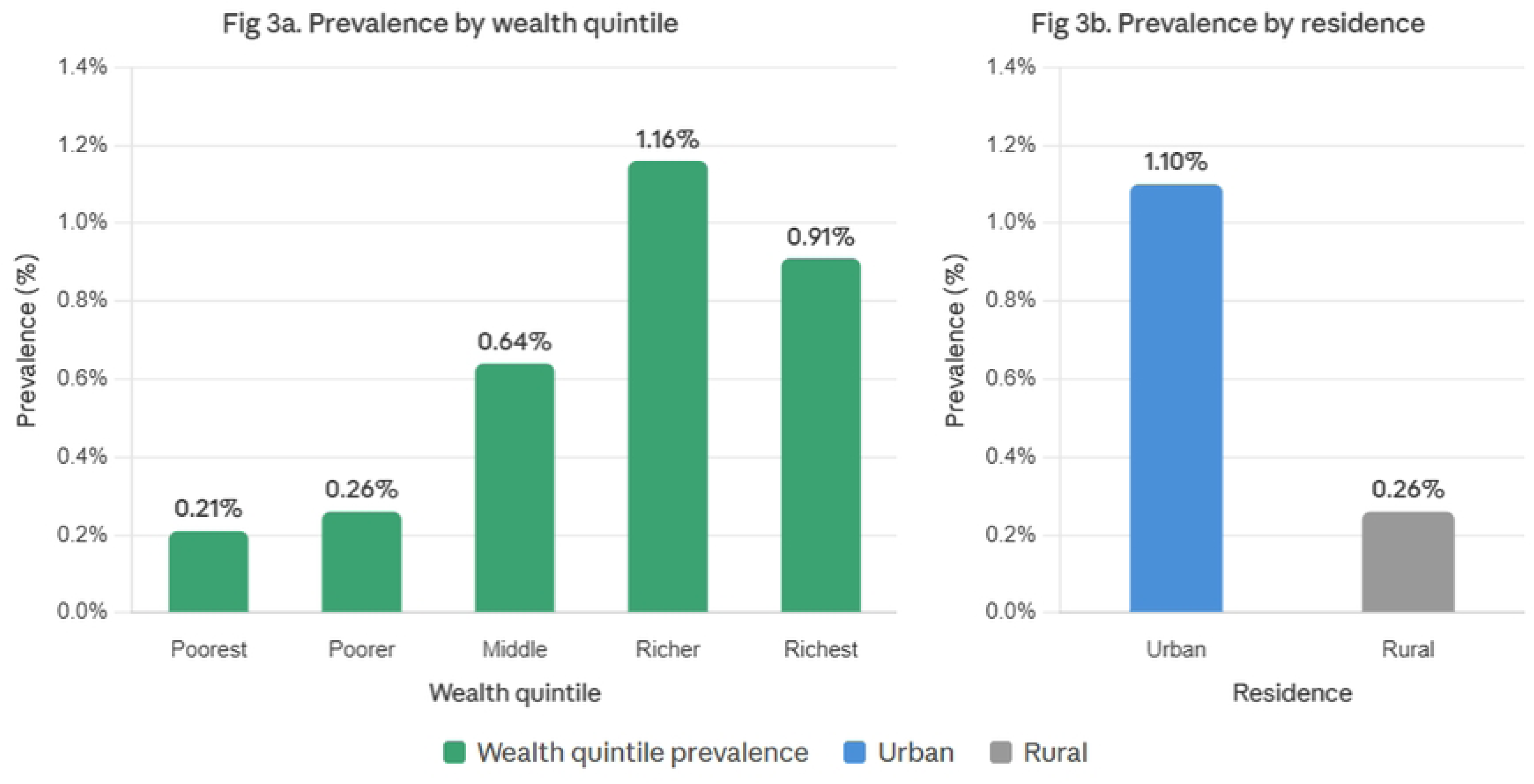
HIV prevalence by (A) household wealth quintile and (B) type of residence among children of HIV-negative mothers, ZDHS 2024.

### Prevalence by child sex

Male children had a weighted HIV prevalence of 0.70% (42 of 7,017), compared with 0.46% (27 of 6,943) among females. Although the point estimate of the odds ratio suggested a modest elevation in risk among males (OR = 1.54, 95% CI: 0.95–2.50), this association did not reach statistical significance (*p* = 0.10).

### Prevalence by child age group

Prevalence was broadly similar across age groups: 0.52% among children aged 2–4 years, 0.50% among those aged 5–9 years, and 0.75% among those aged 10–14 years. The chi-square test indicated no statistically significant variation by age group (*p* = 0.29).

### Prevalence by household wealth

A clear positive gradient in HIV prevalence across wealth quintiles was observed (Table 1; Fig 3A; *p* < 0.001). Prevalence increased from 0.21% in the poorest quintile to 1.16% in the richer quintile, with a slight decline to 0.91% in the richest quintile. Compared with the poorest quintile, the odds of HIV positivity were significantly elevated in the richer quintile (OR = 4.93, 95% CI: 2.19–11.10) and the richest quintile (OR = 4.31, 95% CI: 1.82–10.18).

### Prevalence by mother’s education

HIV prevalence among children showed a rising trend with increasing maternal education: 0.43% among children of mothers with no education, 0.42% for primary education, 0.80% for secondary, and 1.07% for higher education. This association was marginally statistically significant (*p* = 0.04).

## Discussion

### Principal findings

This secondary analysis of the ZDHS 2024 revealed that 0.58% of children aged 2–14 years whose mothers tested HIV-negative were themselves HIV-positive. Three key correlates emerged: geographic province, urban residence, and household wealth. Notably, Copperbelt Province exhibited a prevalence of 2.97%—approximately five times the national average—a finding that demands further investigation. Urban residence conferred a nearly fourfold increase in odds compared with rural residence, and children in wealthier households were paradoxically more likely to be HIV-positive than those in the poorest households.

### Burden in comparative context

The finding of HIV-positive children among women testing HIV-negative is consistent with prior studies from sub-Saharan Africa. Analyses of DHS data from Mozambique, Malawi, and Zimbabwe have reported similar, albeit small, proportions of seropositive children born to seronegative mothers [10,11,13]. The phenomenon has been attributed to several mechanisms, including healthcare-associated transmission through unsafe injections or blood products, and, particularly among older children, sexual abuse [8,9,14].

### Geographic and urban concentration

The strikingly high prevalence in Copperbelt Province is a novel finding that, to our knowledge, has not been reported in previous Zambian studies. The 2024 ZDHS confirmed that Copperbelt remains one of the provinces with the highest adult HIV prevalence nationally at 11.6% [1], and national HIV estimates for 2025 identified Copperbelt as the province with the highest number of new HIV infections [5]. Copperbelt is one of Zambia’s most urbanised and industrialised provinces, with substantial population mobility associated with the mining sector [15,16]. The concentration of cases in this province may reflect a combination of higher community-level viral exposure, greater population density facilitating non-vertical transmission pathways, and increased opportunities for healthcare-associated or other non-MTCT exposures in a high-prevalence setting. Mining communities in particular have been associated with elevated HIV risk due to transactional sex, migration, and limited health service continuity [17].

The urban–rural disparity is consistent with the broader epidemiology of HIV in Zambia, where urban areas have historically borne a disproportionate burden of infection [1,2]. Urban settings may confer higher risk through greater exposure to healthcare settings with variable infection control standards, denser social networks facilitating transmission, and, among older children, increased vulnerability to sexual violence [18].

### Wealth gradient

Perhaps the most counterintuitive finding was the positive association between household wealth and child HIV prevalence. This pattern stands in contrast to the conventional understanding that poverty drives HIV risk through mechanisms such as transactional sex, limited healthcare access, and food insecurity [19]. However, the relationship between wealth and HIV in sub-Saharan Africa is more complex than often assumed. Several studies have documented higher HIV prevalence among wealthier individuals in generalised epidemics, a phenomenon attributed to greater social and sexual networking, higher mobility, and increased access to urban environments where transmission is more intense [20,21]. In the present context, the wealth gradient may also be partially confounded by geography, as wealthier households are disproportionately concentrated in urban and Copperbelt areas where prevalence is highest. Without multivariable adjustment—precluded here by the small case count—it is not possible to determine whether wealth is an independent risk factor or a proxy for geographic and environmental exposures.

### Sex differences and age patterns

Neither child sex nor age group was significantly associated with HIV status, although point estimates suggested a slightly higher prevalence among males and among older children aged 10–14 years. The lack of statistical significance likely reflects the limited power conferred by only 69 positive cases rather than a true absence of effect. The modestly elevated prevalence among older children could be consistent with cumulative exposure over time or, for adolescents, sexual transmission including through abuse. Future studies with larger sample sizes should revisit these associations.

The marginal association with maternal education (*p* = 0.04) followed a gradient similar to that observed for wealth, with higher prevalence among children of more educated mothers. This pattern likely reflects the same urban–wealth confounding described above and is consistent with findings from other DHS-based studies in the region [20].

### Public health implications

These findings have several programmatic implications. First, the concentration of cases in Copperbelt Province suggests the need for province-specific programmatic responses, including strengthened infection prevention and control in healthcare facilities, community-based surveillance for non-MTCT paediatric infections, and targeted paediatric HIV case-finding initiatives in mining and urban communities. Second, the urban–rural disparity underscores the importance of targeting urban health systems for improved paediatric HIV case finding. Third, the paradoxical wealth gradient challenges simplistic assumptions about the social determinants of paediatric HIV and calls for more nuanced, context-specific approaches to risk identification.

At the national level, these results highlight the need for improved surveillance of healthcare-associated HIV transmission, including tracking of injection safety indicators, blood product screening protocols, and facility-level infection prevention and control audits [23]. Given that the mothers of these children tested HIV-negative, interventions should focus on non-vertical transmission pathways: ensuring safe medical practices in paediatric healthcare settings, strengthening child protection systems to prevent and respond to sexual abuse, and conducting epidemiological investigations in provinces with unusually high prevalence to identify specific sources of transmission. These recommendations are particularly urgent in light of ongoing reductions in PEPFAR and Global Fund support, which threaten to weaken the very systems needed to identify and respond to non-MTCT paediatric infections [7].

### Limitations

This study has several limitations. The small number of HIV-positive cases (n = 69) severely constrains statistical power and precision, producing wide confidence intervals and precluding multivariable regression analysis. The cross-sectional study design does not permit the establishment of causal relationships or the sequencing of events. DHS data do not capture information on healthcare exposure history, history of sexual abuse, or other non-MTCT risk factors, limiting the ability to characterise transmission pathways. Finally, the possibility of laboratory error, although minimised by the DHS quality assurance protocol, cannot be entirely excluded. Despite these limitations, the nationally representative design, standardised DBS testing methodology, and large sample size of 13,960 children strengthen the generalisability of these findings.

## Conclusions

Among children aged 2–14 years of HIV-negative mothers in Zambia, HIV prevalence was 0.49% (survey-weighted: 0.58%). The marked geographic concentration in Copperbelt Province, the strong association with urban residence, and the paradoxical positive wealth gradient collectively suggest that non-MTCT transmission pathways play an important role in this population. Healthcare-associated transmission and child sexual abuse represent plausible mechanisms that warrant further investigation. These findings call for enhanced infection prevention and control in healthcare facilities, strengthened child protection systems, targeted surveillance in high-prevalence provinces, and further research employing prospective designs and detailed exposure assessments to elucidate the specific pathways of HIV acquisition in this vulnerable population.

## Data Availability

The datasets analysed in this study are publicly available from The DHS Program data repository (https://dhsprogram.com). Specifically, the 2024 Zambia Demographic and Health Survey HIV Test Recode (ZMAR81FL) and Person Recode (ZMPR81FL) datasets can be accessed upon free registration and approval of a data access request through The DHS Program website.

## Acknowledgments

The authors acknowledge the Zambia Statistics Agency, the Ministry of Health, and ICF for making the ZDHS 2024 data publicly available. The authors also acknowledge all survey participants and field teams who contributed to the 2024 ZDHS.

## Supporting information

**S1 Fig.** Participant flow diagram

**S2 Fig**. HIV prevalence by province among children of HIV-negative mothers, ZDHS 2024

**S3 Fig.** HIV prevalence by (A) household wealth quintile and (B) type of residence among children of HIV-negative mothers, ZDHS 2024.

**S1 Table.** Full provincial breakdown of HIV prevalence with unweighted cell counts among children born to HIV-negative mothers, ZDHS 2024.

**S1 Checklist.** STROBE Statement

**S1 Other.** Human Participants Research Checklist.

**S1 Text.** Python Analysis Code

**Data Review URL**

